# Self-reported symptoms, self-reported viral testing result and seroprevalence of SARS CoV-2 among a community sample in Essex County New Jersey: A brief report

**DOI:** 10.1101/2021.03.02.21252766

**Authors:** Henry F. Raymond, Pratik Datta, Rahul Ukey, Peng Wang, Richard J. Martino, Kristen D. Krause, Corey Rosmarin-DeStefano, Abraham Pinter, Perry N. Halkitis, Maria L. Gennaro

## Abstract

**Background:** SARS-CoV-2, the virus that causes COVID-19, has rapidly spread globally beginning in late 2019. Early areas impacted by this pandemic in the US include Essex County, New Jersey. Beyond understanding the prevalence of active infections and deaths, it is important to understand the true burden of infection in the community, as indicated by seroprevalence of antibodies directed to the virus. Understanding the spectrum of disease is key to the effectiveness of primary prevention and control measures and the design of interventions against transmission of infection.

**Methods:** We utilized venue-based-sampling (VBS), implemented by a community partner, to sample members of the community in Essex County. In VBS the venues are randomized as a proxy for randomizing the attendees of the venues. We asked standard demographic questions, questions about symptoms and PCR testing and previous antibody testing. Participants provide a blood sample collected by finger stick with the Neoteryx Mitra Collection device. Samples were tested using a novel ELISA based approached developed by our team.

**Results:** From September 15, 2020 to December 22, 2020, we conducted 92 randomly selected sampling events where we approached 1349 individuals for screening. Of these, 924 consented and had complete data for analysis. Only 6.5% of the sample reported any COVID-19 like symptoms while 45.9% had sought out a COVID-19 test. In total 13 (1.4%) participants received a positive SARS-CoV-2 PCR test result. While 33 participants (2.6%) sought a SARS-CoV-2 antibody test, only 0.5% of the sample reported a positive antibody result. Testing in this study identified 83 (9.0%) participants positive for SARS-CoV-2 antibodies.

**Conclusion:** We recruited a large sample of the population of Essex County, New Jersey using VBS, electronic surveys, novel sample collection and lab methods. Our findings suggest that the burden of SARS-Cov-2 is slightly more than six times than that suggested by PCR testing. This burden is higher than most estimates obtained through studies of remnant blood samples from hospitals (4.2%), samples from staff at a public-school system (2.9%), and residents of a California county recruited with targeted Facebook ads (1.5%). (9-11) Moreover, with only 6.5% of the sample reporting any COVID-19-like symptoms, our finding suggests that the number of asymptomatic persons may be close to 1.5 times greater than anyone reporting symptoms.

## Introduction

SARS-CoV-2, the virus that causes COVID-19, has rapidly spread globally beginning in late 2019. As of mid-February 2021, 109,659,317 cases of SARS-CoV-2 have been identified worldwide with a corresponding 2,423,307 deaths. At the same time 27,762,686 cases have been identified with a corresponding 488,352 deaths in the US. Early areas impacted by this pandemic in the US centered around the New York metropolitan area and parts of New Jersey. One of these jurisdictions is Essex County, New Jersey.

Essex County is densely populated (6,000 residents per square mile) and diverse (largely African American and Latinx; 62.7%). Moreover, Essex County has large numbers of people living in poverty (15.8%), with moderate income inequality, and over 12% uninsured.(1) Thus, it is likely that a large proportion of the population may not have had the ability to take precautionary measures (e.g., social distancing, self-quarantine) or adequate access to testing and medical care. Indeed, as of mid-February 2021 Essex County ranks second in terms of cases and deaths in New Jersey with 64,287 cases and 2,348 deaths among a population of approximately 795,404 people.(1)

In addition to understanding the prevalence of active infections and deaths, it is important to understand the actual pervasiveness of the burden of infection in the community, as indicated by seroprevalence of antibodies directed to the virus, for a number of reasons. Populations, such as those in Essex County are frequently high-risk for health challenges, and health disparities often go hand in hand with social conditions.(2) In effect, capturing antibody prevalence data helps better understand the cause of spread of infection and how it correlates to the upstream determinants of health and the impact of COVID-19 on communities. Moreover, understanding the spectrum of disease in communities is key to the effectiveness of primary prevention and control measures and the design of rational interventions against transmission of infection moving forward. The total number of infected individuals, those with symptoms, and those with SARS-CoV-2-specific antibodies can help monitor the epidemic and may be helpful for the roll out of immunization programs. With this in mind, we present estimates of COVID-19 symptoms, self-reported results of PCR testing for virus, self-reported antibody results for the virus, and our results of antibody testing in a sample of the community in Essex County, New Jersey.

## Methods

We utilized venue-based-sampling (VBS) to sample members of the community in Essex County. VBS is often employed as an alternative sampling method where simple random sampling is not feasible. In VBS the venues are randomized as a proxy for randomizing the attendees of the venues. For example, VBS is the standard to sample certain populations at high-risk for HIV infection where the population is known to attend identifiable venues. VBS is also the standard sampling method used by CDC for men who have sex with men in the US in National HIV Behavioral Surveillance (NHBS) and is used for other populations internationally.(3-5) North Jersey Community Research Initiative (NJCRI), which has over 20 years’ experience in VBS as the implementing partner in NHBS for New Jersey, performed the VBS sampling for the present study by utilizing their mobile units, which have medical equipment, survey and phlebotomy space, and a private exam room. These units were also outfitted for COVID-19 by adding additional air filtration units and plexi-glass dividers to keep participants and staff safe. We identified venues using Google.com searches using the key words “Supermarkets in Essex County” and “Groceries in Essex County” to search for all potential locations. We identified 205 potential locations. Days and hours of operation were verified by contacting each venue directly. Venues, days, and times of operation were entered in an Excel spreadsheet. To map the venues, we used R 4.0.2 GUI (The R Foundation for Statistical Computing, Austria) to manipulate and analyze the data. Specifically, we used “tidygeocoder” package 1.0.1 to extract coordinates of each store from US Census and OpenStreetMap (OSM) dataset. For each month of sampling approximately 42 venue-day-times were randomly sampled and scheduled for a sampling event. NJCRI’s mobile units and staff used the VBS schedule and travelled to these grocery store sites. At the site, potential participants were systematically approached for a standard two-hour period of time, asked to stop for screening (intercept), screened for eligibility and, if eligible, invited to participate and provide verbal informed consent. Participants who agreed to participate completed a short computer-based survey administered by study staff, using a tablet computer.

The survey included questions on basic self-reported demographics, COVID-19 symptoms (including: fever, dry cough, shortness of breath, chills, headache, sore throat, muscle pain/ aches, new loss of taste or smell, GI issues, neurologic symptoms) since January 2020, seeking viral PCR testing, PCR results, seeking antibody testing and antibody results.

Participants were then asked to provide a blood sample collected by finger stick with the Neoteryx Mitra Collection device to use for SARS-CoV-2 antibody testing.(6) Dried sticks were preserved by the NJCRI collection team or at the testing laboratory at room temperature for up to a week, according to manufacturer’s instructions. At the time of testing, dried blood from the Mitra device was eluted overnight at 4C, based on published protocols.(6) The eluate was diluted with 2% non-fat dry milk to1:80 and utilized for detection of IgG antibodies to recombinant receptor binding domain (RBD) of the SARS-CoV-2 Spike (100 ng/well), utilizing standard enzyme-linked immunosorbent assay (ELISA) protocols.(7, 8) To interpret the ELISA data, we calculated a cut off of OD405 = 0.3 (mean +3 standard deviations) by testing finger-stick eluates from 50 COVID-19 PCR-negative subjects.

Participants were given a result-return card to enable them to call in to the study team for their antibody results a week after sample receipt by the laboratory. Participants received $25 in cash for their participation. This study had ethical approval from the Rutgers IRB. Survey data and lab data were merged in SAS (v9.4). All analyses were conducted in SAS (v9.4) using tabulations.

## Results

From September 15, 2020 to December 22, 2020, we conducted 92 randomly selected sampling events from geographically diverse locations in Essex County where we systematically approached patrons for screening (Figure 1.). We approached 1349 individuals for screening. Of these, 928 (68.8%) stopped and agreed to be screened, 928 (100%) were eligible and agreed to participate, and 924 (99.6%) consented and had complete survey and laboratory data for analysis.

**Figure 1.**
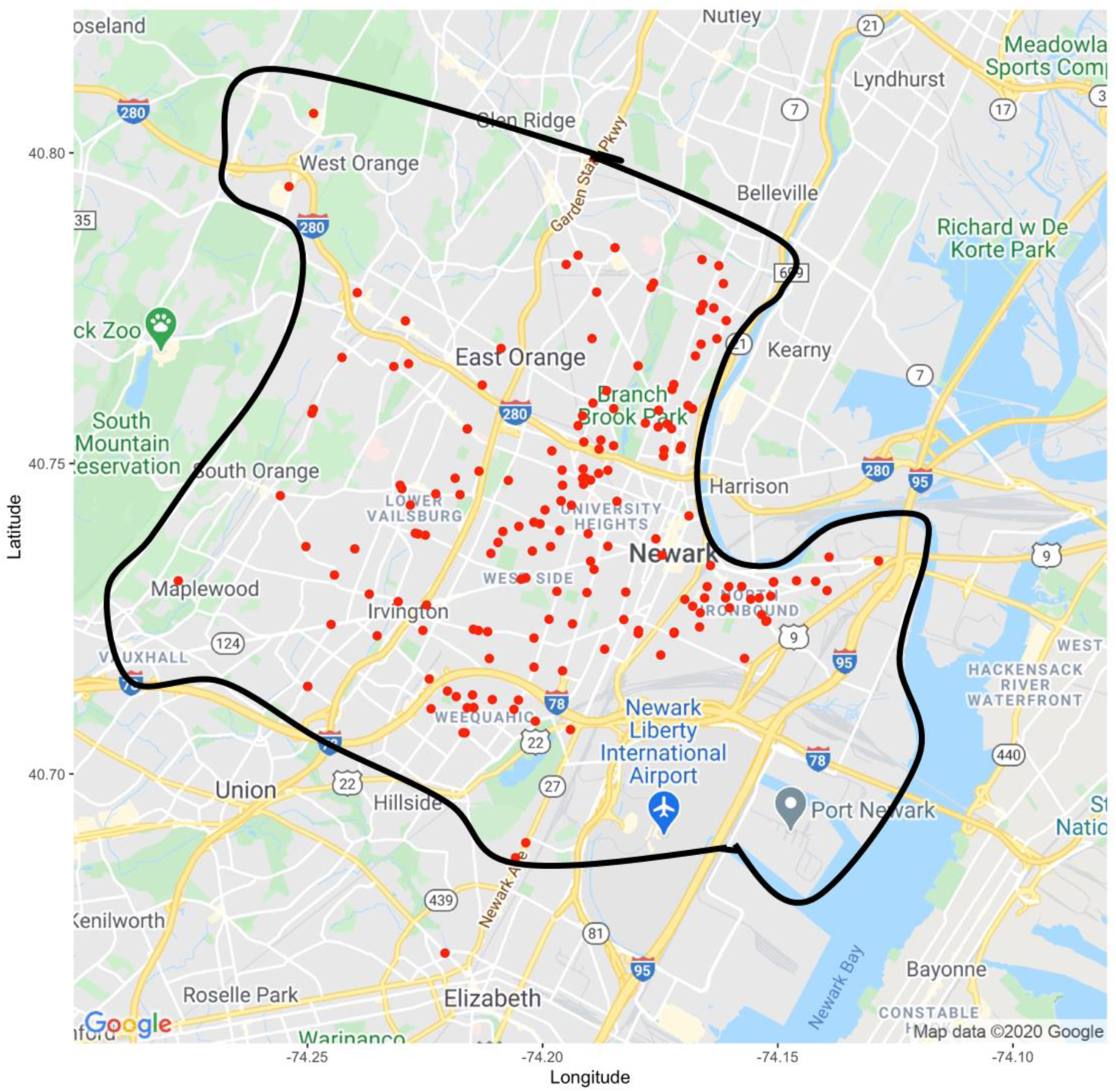
Spatial distribution of venues in Essex County, NJ: study of SARS-Cov-2 seroprevalence, 2020.

Our sample population was mostly over 39 years of age (n=593, 64.2%) with a mean age of 44.7 years, mostly male at birth (n=584, 63.2%), predominantly Black (n=656, 71.0%) and Latino (n=194, 21.0%). A majority had a high school diploma or some college education (n=652, 70.5%). Only 343 (37.1%) were employed full or part-time, while 398 (43.1%) reported being unemployed. Personal annual income reported by participants was low, with almost half, 442 (47.8%), reporting a personal annual income of less than $15,000 while another 300 (32.5%) reported an income of between $15,000 and $35,000. Homelessness was common in our sample, with 251 (27.2%) reporting being homeless sometime in the 12 months preceding the survey and 181 (19.6%) reporting being homeless at the time of the study. A majority of the sample reported living in Newark (n=704, 76.2%). Finally, 77.9% (n=720) of the sample reported having some sort of health insurance at the time of the study. (Table 1.)

**Table 1.**
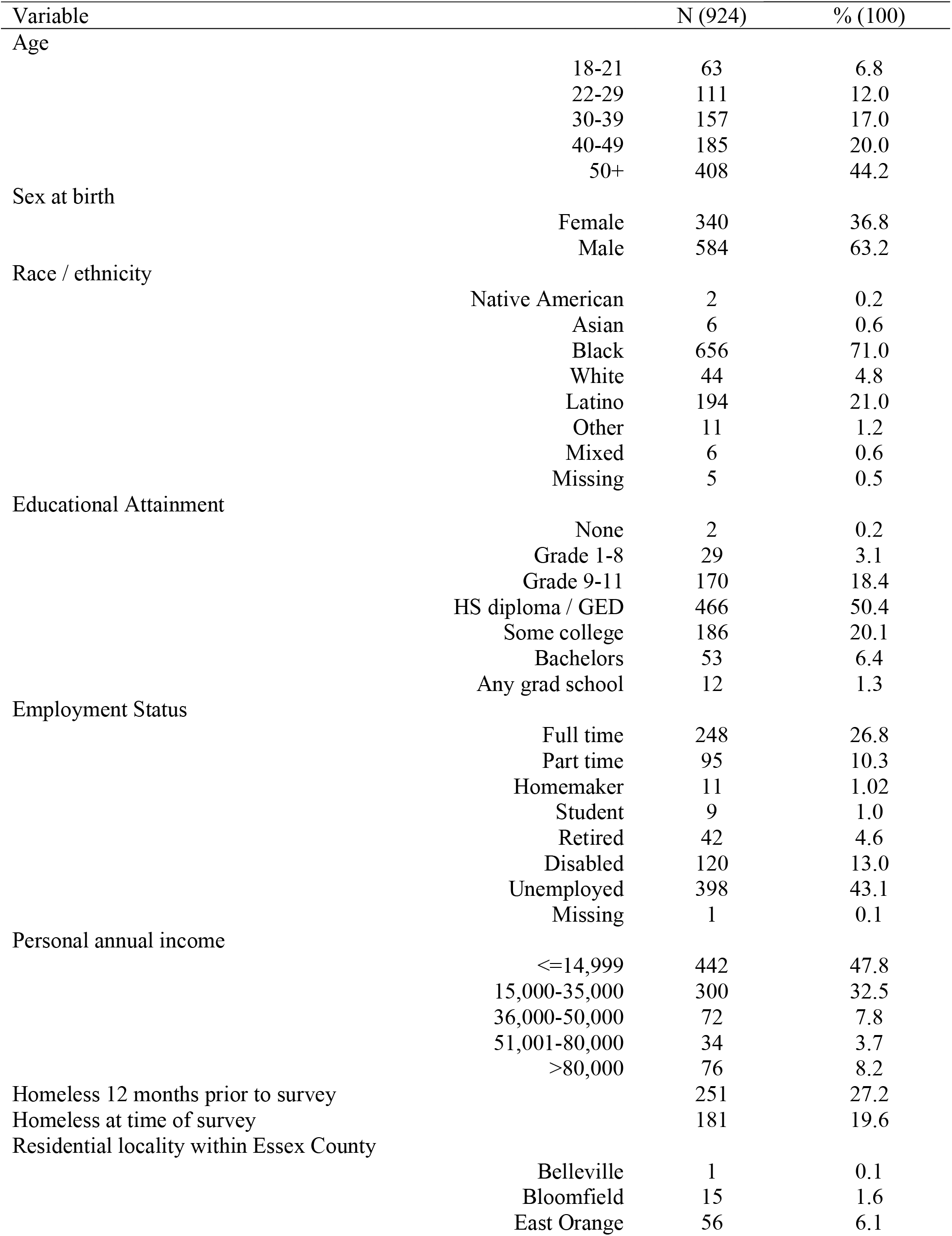
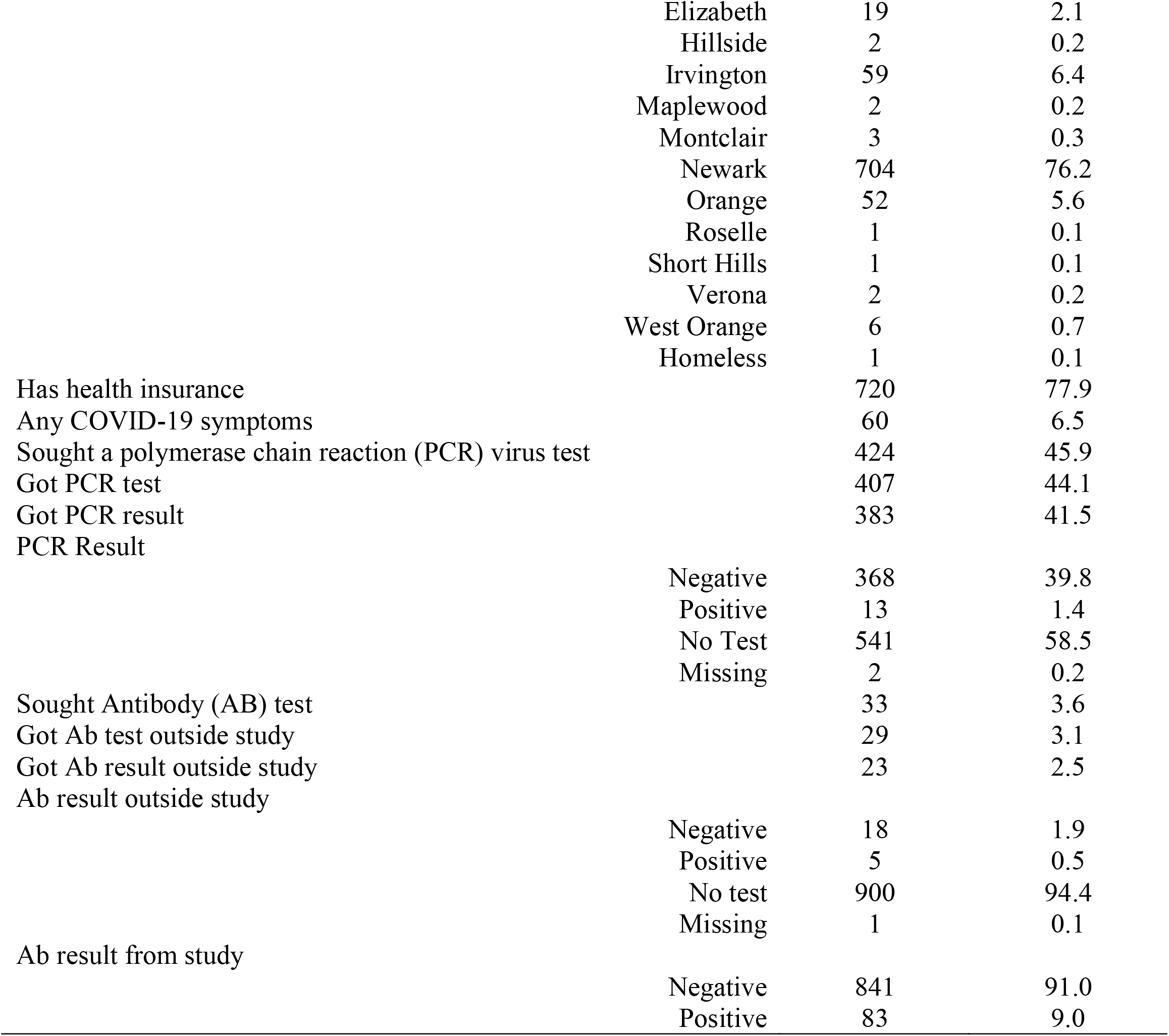
Demographics, symptoms, testing seeking indicators and prevalence of SARS-CoV-2 antibody prevalence in a community sample, Essex County, NJ, 2020.

| Variable | N (924) | % (100) |
| --- | --- | --- |
| Age |  |  |
| 18-21 | 63 | 6.8 |
| 22-29 | 111 | 12.0 |
| 30-39 | 157 | 17.0 |
| 40-49 | 185 | 20.0 |
| 50+ | 408 | 44.2 |
| Sex at birth |  |  |
| Female | 340 | 36.8 |
| Male | 584 | 63.2 |
| Race / ethnicity |  |  |
| Native American | 2 | 0.2 |
| Asian | 6 | 0.6 |
| Black | 656 | 71.0 |
| White | 44 | 4.8 |
| Latino | 194 | 21.0 |
| Other | 11 | 1.2 |
| Mixed | 6 | 0.6 |
| Missing | 5 | 0.5 |
| Educational Attainment |  |  |
| None | 2 | 0.2 |
| Grade 1-8 | 29 | 3.1 |
| Grade 9-11 | 170 | 18.4 |
| HS diploma / GED | 466 | 50.4 |
| Some college | 186 | 20.1 |
| Bachelors | 53 | 6.4 |
| Any grad school | 12 | 1.3 |
| Employment Status |  |  |
| Full time | 248 | 26.8 |
| Part time | 95 | 10.3 |
| Homemaker | 11 | 1.02 |
| Student | 9 | 1.0 |
| Retired | 42 | 4.6 |
| Disabled | 120 | 13.0 |
| Unemployed | 398 | 43.1 |
| Missing | 1 | 0.1 |
| Personal annual income |  |  |
| <=14,999 | 442 | 47.8 |
| 15,000-35,000 | 300 | 32.5 |
| 36,000-50,000 | 72 | 7.8 |
| 51,001-80,000 | 34 | 3.7 |
| >80,000 | 76 | 8.2 |
| Homeless 12 months prior to survey | 251 | 27.2 |
| Homeless at time of survey | 181 | 19.6 |
| Residential locality within Essex County |  |  |
| Belleville | 1 | 0.1 |
| Bloomfield | 15 | 1.6 |
| East Orange | 56 | 6.1 |

|  |  |  |  |
| --- | --- | --- | --- |
|  | Elizabeth | 19 | 2.1 |
|  | Hillside | 2 | 0.2 |
|  | Irvington | 59 | 6.4 |
|  | Maplewood | 2 | 0.2 |
|  | Montclair | 3 | 0.3 |
|  | Newark | 704 | 76.2 |
|  | Orange | 52 | 5.6 |
|  | Roselle | 1 | 0.1 |
|  | Short Hills | 1 | 0.1 |
|  | Verona | 2 | 0.2 |
|  | West Orange | 6 | 0.7 |
|  | Homeless | 1 | 0.1 |
| Has health insurance |  | 720 | 77.9 |
| Any COVID-19 symptoms |  | 60 | 6.5 |
| Sought a polymerase chain reaction (PCR) virus test |  | 424 | 45.9 |
| Got PCR test |  | 407 | 44.1 |
| Got PCR result |  | 383 | 41.5 |
| PCR Result |  |  |  |
|  | Negative | 368 | 39.8 |
|  | Positive | 13 | 1.4 |
|  | No Test | 541 | 58.5 |
|  | Missing | 2 | 0.2 |
| Sought Antibody (AB) test |  | 33 | 3.6 |
| Got Ab test outside study |  | 29 | 3.1 |
| Got Ab result outside study |  | 23 | 2.5 |
| Ab result outside study |  |  |  |
|  | Negative | 18 | 1.9 |
|  | Positive | 5 | 0.5 |
|  | No test | 900 | 94.4 |
|  | Missing | 1 | 0.1 |
| Ab result from study |  |  |  |
|  | Negative | 841 | 91.0 |
|  | Positive | 83 | 9.0 |

Only 6.5% of the sample reported any COVID-19 like symptoms since January, 2020 while 45.9% reported having sought out a COVID-19 test. In total 13 (1.4%) participants reported receiving a positive SARS-CoV-2 PCR test result. While 33 participants (2.6%) sought a SARS-CoV-2 antibody test outside of the study, only 0.5% of the sample reported a positive antibody result. Testing conducted in this study identified 83 (9.0%) participants as positive for SARS-CoV-2 antibodies. (Figure 2.)

**Figure 2.**
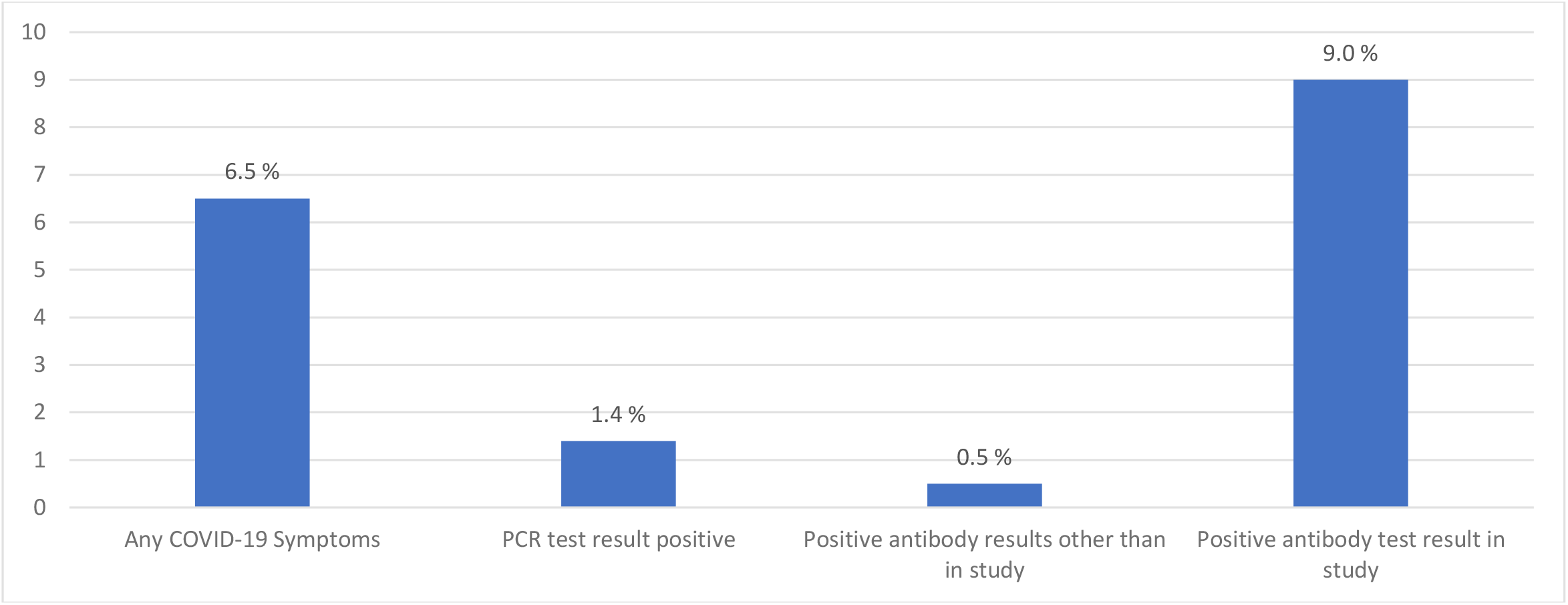
COVID-19 and SARS-CoV-2 indicators among a community sample, Essex County, NJ, 2020.

## Discussion

We successfully recruited a large sample of the population of Essex County, New Jersey for this seroprevalence study. Our sample was predominantly Black and Latino and residing in Newark. Importantly, our findings suggest that the true burden of SARS-Cov-2 and the pool of persons potentially spreading the virus is slightly more than six times than that suggested by PCR testing. This burden from our robustly sampled community-based study, is higher than most estimates obtained through studies of remnant blood samples from hospitals (4.2% antibody prevalence), samples from staff at a public-school system (2.9% antibody prevalence), and residents of a California county recruited with targeted Facebook ads (1.5% antibody prevalence). These differences underscore the importance of conducting robust community-based sampling to determine the true burden of disease. (9-11) Moreover, with only 6.5% of the sample reporting any COVID-19-like symptoms, our finding suggests that the number of asymptomatic persons may be close to 1.5 times greater than anyone reporting symptoms (it is noted however that the symptoms listed might include other conditions).

Our selection of Spike RBD as viral antigen for antibody detection has at least two advantages over commercial serological assays for SARS-CoV-2 infection, which are mostly based on detecting antibodies against the viral Nucleocapsid (N) protein. One is that circulating RBD-specific antibodies may last longer than N-specific antibodies (13) and are therefore more reliable markers of infection for seroprevalence studies (at least in non-vaccinated subjects). In addition, RBD is the portion of the SARS-CoV-2 Spike protein that establishes direct contact with the receptor on the host cells (14) and, consequently, is the main target of neutralizing antibodies (15), which are protective. Thus, our results indirectly suggest that at least 9.0% of the study population likely expresses some protective immunity.

Our study has limitations. VBS may not reach individuals who do not do their own shopping or live in settings where food is provided, thus introducing some form of sampling bias. Secondly, while Essex County is predominantly populated by Black / Latino individuals, our study oversampled these groups. This may be due to lower enthusiasm for participation among other race/ethnicity groups that may not be as altruistic or interested in the incentive provided by the study.

Despite these limitations, we were able to sample a large number of individuals and estimate SARS-CoV-2 antibody prevalence in a county highly impacted by the current pandemic. Our findings underscore the importance of conducting seroprevalence studies in addition to case reporting to understand the true magnitude of SARS-CoV-2 impact on communities and plan for mitigation strategies to reduce transmission.

## Supporting information

STROBE Checklist

## Data Availability

Data may be available upon request.

